# Cross-Cultural Validation of Acculturation Measures: Expanding the East Asian Acculturation Framework for Global Applicability

**DOI:** 10.1101/2024.09.01.24312903

**Authors:** Erhabor Sunday Idemudia, Constance Karing, Lawrence Ejike Ugwu

## Abstract

In a globalised world, understanding acculturation—the process by which individuals adapt to new cultural environments—is crucial, especially in multicultural societies. The East Asian Acculturation Measure (EAAM), rooted in Berry’s acculturation model, has been extensively used to assess acculturation strategies among East Asian populations in the United States. However, its application to other cultural groups remains limited due to its specificity. This study aims to adapt and validate the EAAM for broader applicability across diverse cultural contexts, particularly among populations outside the East Asian demographic.

The study involved 819 international students in Germany and South Africa from 112 countries. It was conducted in two phases: the first included 490 and 329 in the second. The psychometric properties of the EAAM were evaluated through Confirmatory Factor Analysis (CFA) and Exploratory Factor Analysis (EFA). Initial CFA results showed that the original four-factor model did not adequately fit the diverse sample, prompting further EFA, which revealed a more suitable five-factor structure. This new structure termed the Shortened Adapted Acculturation Scale (SAAS), comprises five dimensions: Social Disconnection, Cultural Adaptation, Social Perception, Interpersonal Comfort, and Language Integration. The SAAS demonstrated high internal consistency and reliability across all factors, with significant evidence of measurement invariance across genders.

The findings emphasise the importance of culturally adapting psychological measures to ensure validity and reliability across diverse populations. The SAAS offers a robust tool for assessing acculturation beyond East Asian contexts, providing valuable insights into the complexities of cultural adaptation. The study’s outcomes contribute significantly to the field of cross-cultural psychology and highlight the need for continued research on acculturation processes in an increasingly diverse world.

## Introduction

In today’s globalised world, understanding the processes of acculturation—how individuals adapt to new cultural environments—is increasingly critical. Accurate measurement of acculturation is essential for capturing the complexities of cultural adaptation, particularly in multicultural societies where diverse cultural groups interact. These measurements are crucial for academic research and developing culturally sensitive interventions that support individuals’ psychological and social well-being across different cultural contexts (Sousa & Rojjanasrirat, 2011). Recent advancements in acculturation research emphasise the importance of reliable and valid measures applicable across various cultural settings, ensuring that they effectively capture the complexities of cultural adaptation (Park et al., 2021).

The East Asian Acculturation Measure (EAAM) was initially developed to assess the acculturation patterns of East Asian immigrants in the United States, based on Berry’s acculturation model, which categorises acculturation strategies into assimilation, separation, integration, and marginalisation (Berry, 2001). While the EAAM has been a valuable tool in understanding acculturation among East Asian populations, its design reflects cultural and social dynamics unique to East Asians, limiting its effectiveness when applied to other cultural groups, such as those in South Asia, Africa, or Latin America (Chung et al., 2004). This cultural specificity presents significant challenges for researchers and practitioners using the EAAM in more diverse populations (Okazaki, 1998; Ramírez et al., 2005).

To overcome these limitations, there is a clear need to adapt and validate the EAAM for broader cultural applicability. Such adaptation ensures that the scale accurately captures the acculturation processes of individuals from diverse backgrounds, considering the distinct social, linguistic, and cultural dynamics they encounter. Recent efforts to adapt acculturation measures, such as the development of the Acculturation and Resilience Scale (AARS), underline the importance of incorporating cultural specificity to enhance the validity and reliability of these tools across different cultural groups (Khawaja et al., 2014). Moreover, the adaptation of scales such as the Brief Sociocultural Adaptation Scale (BSAS) and the Brief Psychological Adaptation Scale (BPAS) for use in different linguistic and cultural contexts demonstrates the necessity of modifying existing measures to reflect the diverse experiences of global populations (Ohki & Vachkov, 2022).

This follow-up study is essential in evolving the EAAM into a versatile and culturally sensitive tool that can be reliably used across diverse cultural contexts. By adapting the scale, this research aims to equip researchers and practitioners with a robust instrument for assessing acculturation, thereby enhancing our understanding of how different cultural groups navigate the complexities of cultural adaptation. The study will focus on adapting the EAAM for use in non-East Asian cultural contexts, ensuring it accurately captures acculturation processes in diverse groups such as South Asia, Africa, and Latin America. Additionally, it will evaluate the psychometric properties of the adapted scale across various cultural contexts to establish measurement invariance across genders, ensuring that the scale measures acculturation consistently and comparably across different groups (Zea et al., 2003). This is crucial as recent studies have highlighted the importance of ensuring that acculturation measures are reliable and valid across different cultural and linguistic contexts (Demes & Geeraert, 2014).

Furthermore, the study will modify scale items to increase cultural sensitivity and specificity, addressing cultural diversity that may affect the accuracy of acculturation measurement by integrating culturally relevant behaviours, attitudes, and experiences into the scale (Cruz et al., 2000). The research will also investigate the relationship between acculturation strategies and stress related to acculturation in various cultural settings to provide insights into how different acculturation strategies impact well-being across different cultural contexts (Vedder et al., 2007). Recent findings suggest how employed acculturation strategies can significantly influence psychological outcomes, making this investigation particularly relevant (Park et al., 2021).

By addressing these objectives, this study will contribute significantly to the field of acculturation research, offering a robust tool for assessing acculturation across diverse populations and enhancing our global understanding of cultural adaptation processes. The outcomes of this study can potentially revolutionise how we approach acculturation, providing valuable insights crucial for developing effective cross-cultural interventions and advancing the theoretical understanding of acculturation.

## Method

### Study 1: Testing of the Psychometric model of the EAAM

#### Participants

Study 1 participants comprised 490 university students from 112 nationalities in Germany and South Africa, whose ages ranged between 18 and 54 (mean age = 26.07 years; SD = 4.38). Two hundred ninety-seven (60.7%) were males, and one hundred ninety-two (39.2%) were females.

### Measures

#### Instrument Section for the East Asian Acculturation Measure (EAAM)

The East Asian Acculturation Measure (EAAM) is a 29-item self-report measure by Berry (2001) to measure the acculturation patterns of participants. The EAAM is a multidimensional aspect of acculturation [assimilation (8 items), separation (7 items), integration (5), and marginalisation (9 items)] among East Asian immigrants. Each item in the EAAM is rated on a 7-point Likert-type scale ranging from 1 (strongly disagree) to 7 (strongly agree). The total score for each subscale is calculated by summing the relevant item scores, with some items reverse-scored as appropriate.

### Procedure

The study adopted a cross-sectional design targeting international students at German universities to assess acculturative strategies. Eligible participants included international students currently enrolled in German universities, aged 18 or older, who had lived in Germany for at least six months. Recruitment was conducted via university email lists, social media, and campus posters, from 31 July 2023 to 30 March, 2024. Participants accessed the survey through a QR code or direct link. The survey, hosted on SoSci Survey and expected to take 15 to 20 minutes, collected data on demographics and acculturative strategies.

Informed consent was obtained electronically (written), and participants were assured confidentiality and their right to withdraw. The Friedrich Schiller University Jena Ethics Committee (FSV 23/049) and North-West University BaSSREC (NWU-01085-22-S7-01) granted ethical approval, and the study adhered to strict ethical guidelines. Data were anonymised, and the study was classified as low-risk. Measures were implemented to address any potential psychological discomfort, including providing counselling resources if needed.

### Data Analysis

Confirmatory factor analysis (CFA) was used for data analysis. We tested the original four-factor structural model. A large class of omnibus tests for overall fit was evaluated with multiple indices, as recommended by Hu and Bentler (1998). The fit of CFA models was determined with chi-square, the critical ratio (X2/df; Bollen, 1989), the root mean squared error of approximation (RMSEA), the root mean square residual (RMR), the standardised root mean squared residual (SRMR), the comparative fit index (CFI), and the goodness of fit index (GFI). However, different cut-off scores have been used to determine how well a model fits the data, and RMSEA of ≤0.06 is generally assumed to reflect an excellent fit to the data (Hu & Bentler, 1999), with scores between 0.06 and 0.08 indicating acceptable fits (Siedlecki, 2007). The RMR and the SRMR are particularly sensitive to misspecified factor covariance. Factor covariance. The RMR should be less than 0.05 to show a good fit, and SRMR of <0.08 indicates the discrepancy between the data and the hypothesised model. Larger values indicate better fit, and CFI and GFI values of 0.90 or larger generally indicate acceptable model fit (Hu & Bentler, 1999). The model fit of the CFA was estimated using AMOS Graphics (version 29).

The analysis showed that the four-factor model did not yield a good fit. An exploratory factor analysis was therefore further conducted to explore possible alternative models. Based on the data we obtained from participants in Study 1, we tested the factor structure to ascertain the EAAM’s underlying dimensionality. We used principal axis factoring (PAF) with the items’ oblique rotation to adapt to the EAAM. To determine its measurement model, AMOS version 29 was used (Brannick, 1995; Hurley et al., 1997; Williams, 1995). A collection of tests for overall fit was evaluated with multiple indices, as recommended by Hu and Bentler (1998).

## Result

A Confirmatory Factor Analysis (CFA) was conducted to test the four factors of the EAAM. The 4-factor model produced a chi-square value of χ^2^(224) = 1371.74, p < .001. The fit indices were: Standardized Root Mean Square Residual (SRMR) = .06, Goodness of Fit Index (GFI) = .85, Normed Fit Index (NFI) = .87, Non-Normed Fit Index (NNFI) = .88, Comparative Fit Index (CFI) = .89, and Root Mean Square Error of Approximation (RMSEA) = .082. All the fit indices like SRMR, NFI, and CFI were below the acceptable ranges of .90 and above, and the RMSEA exceeds the commonly accepted threshold of 0.08, indicating that the model does not adequately fit the data (see Table 1). Therefore, an exploratory factor analysis was conducted to identify possible factors that fit the data.

**Table 1.** The confirmatory factor analysis of the EAAM.

| Model description | chi-square | DF | SRMR | GFI | NFI | NNFI | CFI | RMSEA |
| --- | --- | --- | --- | --- | --- | --- | --- | --- |
| 4-FACTOR | 1371.74 | 224 | .06 | .85 | .87 | .88 | .89 | .082 |

Root Mean Squared Error of Approximation (RMSEA), the root mean square residual (RMR), the standardised root mean squared residual (SRMR), the Comparative Fit Index (CFI), and the Goodness of Fit Index (GFI), Normed Fit Index (NFI), Non-Normed Fit Index (NNFI)

Principal Axis Factoring (PAF) using the original 29 items. The result of the PAF revealed five factors, with 18 items loading on five factors. The other eleven items had lower loadings or cross-loaded on two or more factors and were removed. This process helped in reducing item redundancy. The criteria for loading is a minimum of 0.3 on only one factor. The PAF generated five factors explaining 51.88% of the variance for the complete set of variables. An analysis of the Kaiser–Meyer Olkin (KMO) test of sampling adequacy suggested that the calibration sample was suitable for PAF (KMO = 0.86) with the Bartlett’s Test of Sphericity significant (χ2 = 8600.87; df = 406; p < 0.001). Four items loaded on factor one explained 20.84% of the total variance, four items loaded on factor two explained 14.78% of the variance, factor three had four items too, which explained 7.12% variance, the fourth factor with three items explained 5.87%, and fifth factor with three items explained 3.96% of the total variance. The scree plot further reveals the analysis components’ breaking point. The eigenvalue is plotted against the number of components included in the analysis. Figure 1 shows that the eigenvalue was above 1.00 only among the first five factors.

**Figure 1.**
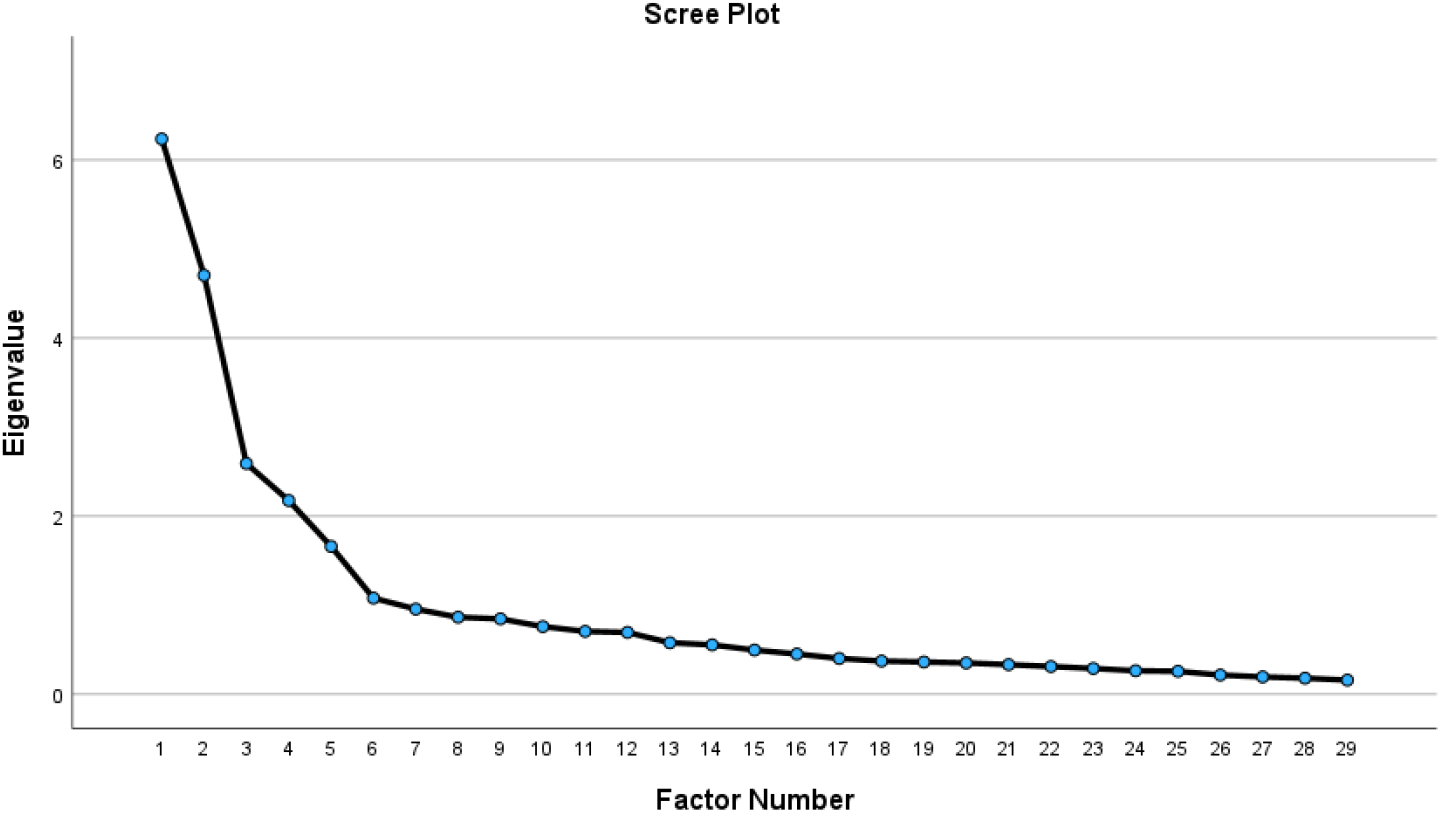
Scree plot for the Five-factor SAAS

## Method

### Study 2: validation and measurement invariance SAAS

#### Participants

Study 2 participants comprised 329 university students from 112 nationalities in Germany and South Africa, aged between 16 and 54 (mean age = 24.04 years; SD = 6.08). One hundred and sixty-six (50.5%) were females, and one hundred and sixty-two (49.5%) were males.

### Measures

#### Instrument Section for the Shortened Adapted Acculturative Strategy (SAAS)

The SAAS is an 18-item self-report measure adapted from the EAAM by Berry (2001) that measures the acculturative strategy of participants. The SAAS is a multidimensional aspect of acculturation [Social Disconnection/social alienation/marginalisation (4-items), Cultural adaptation/social integration (4-items), Cross-cultural social dynamic/cultural affiliation/social perception (4-items), Relationship Preference/interpersonal comfort (3-items) and Cultural expression & Language proficiency/cultural identity & Language dynamic (3-items)] among a general population. The items are rated on a 7-point Likert-type scale ranging from 1 (strongly disagree) to 7 (strongly agree). Examples of items modified: “I get along better with [*country of residence*] than people from my country,” and “Most of my friends at work/school are [*country of residence*].” The total score for each subscale is calculated by summing the relevant item scores.

#### Acculturative Stress Scale for International Students (ASSIS)

The Acculturative Stress Scale for International Students (ASSIS) is a 36-item self-report scale developed by Sandhu and Asrabadi (1994). It was utilised to measure the acculturative stress experienced by international students. It was explicitly designed to assess the psychological challenges that international students face as they adjust to a new cultural environment. The ASSIS addresses a broad range of stressors that contribute to acculturative stress, making it a comprehensive tool for understanding the unique experiences of this population. The ASSIS comprised six primary factors representing different dimensions of acculturative stress [Perceived Discrimination (8 items), Homesickness (4 items), Perceived Hate (5 items), Fear (4 items), Stress Due to Change/Culture Shock (3 items), Guilt (2 items). Each item is rated on a 5-point Likert-type scale ranging from 1 (strongly disagree) to 5 (strongly agree), with higher scores indicating greater levels of acculturative stress.

### Procedure

A cross-sectional design was also adopted for the second study. It focused on international students at South African universities to assess SAAS and acculturative stress. All participants were international students enrolled in South African universities, aged 18 or older, and residents of South Africa for at least six months. Participants were purposively sampled.

Recruitment was conducted through university email lists, social media, and campus posters, with the survey accessible via a QR code or direct link. The survey, conducted on SoSci Survey and expected to take 20 to 25 minutes, gathered data on demographics, acculturative strategies, and stress. Informed consent was obtained electronically, ensuring participants’ confidentiality and their right to withdraw. The study received ethical approval, as stated in study one.

### Data Analysis

We tested the new SAAS five-factor structural model. The fit of CFA models was determined with chi-square, the critical ratio (X2/df; Bollen, 1989), the root mean squared error of approximation (RMSEA), the root mean square residual (RMR), the standardised root mean squared residual (SRMR), the comparative fit index (CFI), and the goodness of fit index (GFI). However, different cut-off scores have been used to determine how well a model fits the data, and RMSEA of ≤0.06 is generally assumed to reflect an excellent fit to the data (Hu & Bentler, 1999), with scores between 0.06 and 0.08 indicating acceptable fits (Siedlecki, 2007). The RMR and the SRMR are particularly sensitive to misspecified factor covariance. Factor covariance. The RMR should be less than 0.05 to show a good fit, and SRMR of <0.08 indicates the discrepancy between the data and the hypothesised model. Larger values indicate better fit, and CFI and GFI values of 0.90 or larger generally indicate acceptable model fit (Hu & Bentler, 1999). The model fit of the CFA was estimated using AMOS Graphics (version 29).

#### Multiple-Group CFA of Invariance Across Gender

Measurement invariance examines whether the assessment of latent constructs is consistent across different groups, as discussed by Cheung and Rensvold (2002) and Kline (2015). This process involves evaluating three critical levels of invariance: configural, metric, and scalar. Configural invariance tests whether the same factor structure is applicable across groups, indicating that the observed variables represent the same pattern of latent constructs in different populations. If configural invariance is supported, it suggests that the basic model holds across groups. However, it does not imply that the relationships between latent constructs and observed variables are identical across those groups (Abrams et al., 2013; Vandenberg & Lance, 2000).

Metric invariance is examined next to ensure the constructs have the same meaning across groups. This level of invariance assesses whether the factor loadings are equivalent, meaning that the constructs are interpreted similarly across different populations. Without metric invariance, comparing latent means across groups is invalid because the constructs may be understood differently (Kline, 2015; Vandenberg & Lance, 2000).

Finally, scalar invariance is tested to determine whether the intercepts of the observed variables are consistent across groups. This is necessary for comparing latent means, as it ensures that the groups have the same baseline level for the latent constructs. If scalar invariance holds, it suggests that differences in means reflect true differences in the latent constructs rather than differences in measurement (Vandenberg & Lance, 2000). The results of these tests for measurement invariance are presented in Table 4.

## Result

### Confirmatory Factor analysis of the SAAS five-factor model

A five-dimensional model of the scale was tested. Each item was constrained to load on the hypothesised dimension. The factor analysis of the data yielded goodness-of-fit indices that support the five-dimensional structure of the adapted scale. The Chi-square goodness-of-fit was significant, with χ2 = 638.95; df = 260; p = 0.001. Furthermore, the other fit indices were all within acceptable limits, with GFI = 0.90, CFI = 0.92, NNFI = 0.91, and RMSEA = 0.061. Recent guidelines suggest the use of the Non-Normed Fit Index (NNFI or TLI), Comparative Fit Index (CFI), and Root Mean Square Error of Approximation (RMSEA) for evaluating model fit. Current standards recommend that NNFI and CFI values above 0.95 indicate a good model fit, and RMSEA values below 0.06 are considered acceptable (Kline, 2015; Hair et al., 2020; Hu & Bentler, 1999). Thus, we considered our model as acceptable. We examined the internal consistency indices of the dimensions (see Figure 2 and Table 2).

**Table 2.** Confirmatory factor analysis of the Five-subscales of SAAS.

| Model description | chi-square | DF | SRMR | GFI | NFI | NNFI | CFI | RMSEA |
| --- | --- | --- | --- | --- | --- | --- | --- | --- |
| 5-FACTOR | 638.95 | 260 | .17 | .86 | .90 | .91 | .92 | .061 |

Note: Root Mean Squared Error of Approximation (RMSEA), the root mean square residual (RMR), the standardised root mean squared residual (SRMR), the Comparative Fit Index (CFI), and the Goodness of Fit Index (GFI), Normed Fit Index (NFI), Non-Normed Fit Index (NNFI)

**Figure 2.**
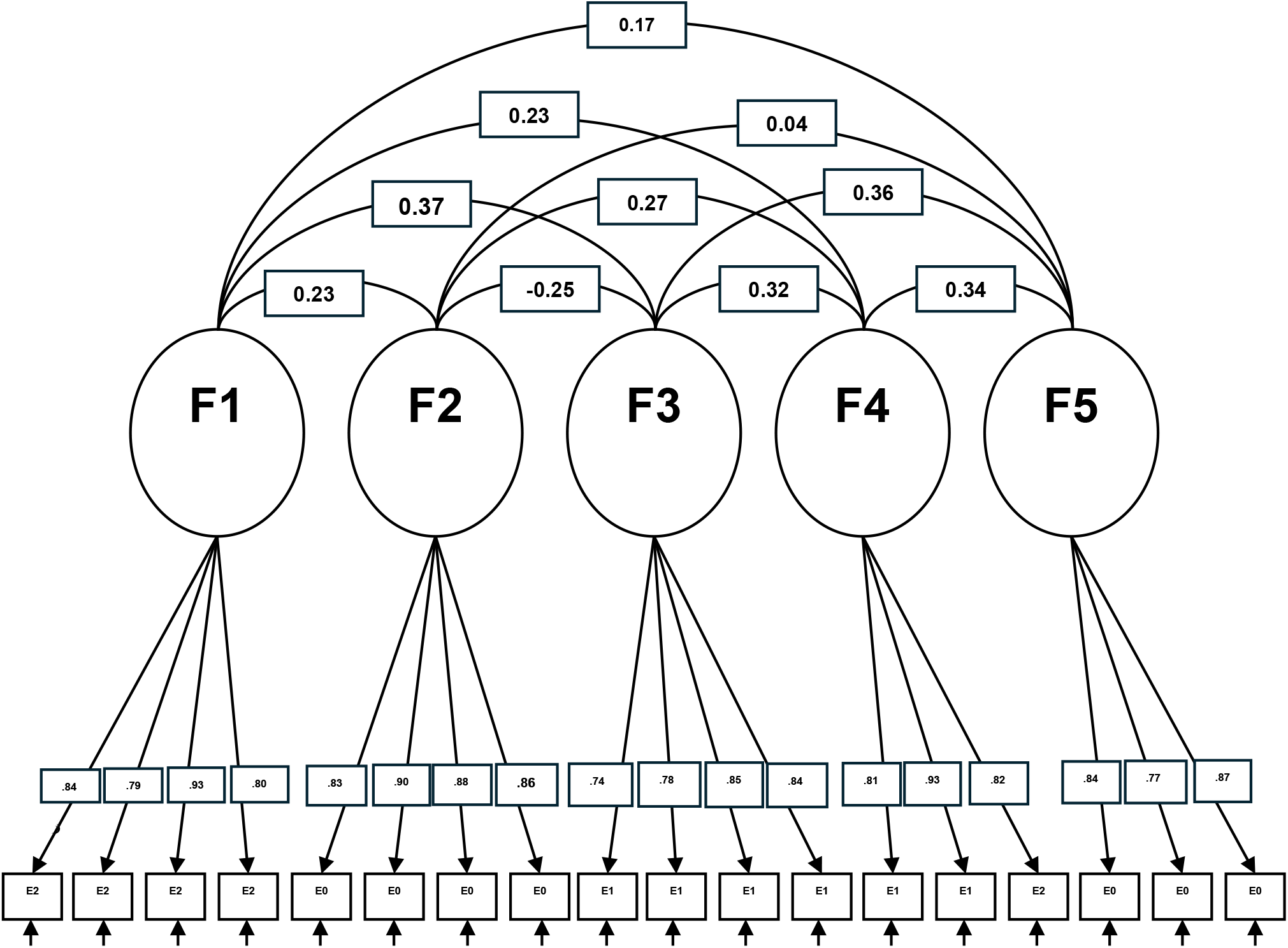
Five-dimensional model for the five subscales of the SAAS.

A measurement model evaluating the scale’s psychometric properties revealed a five-factor structure (see Table 3). The analysis assessed each factor’s internal consistency, reliability, and validity.

**Table 3.**
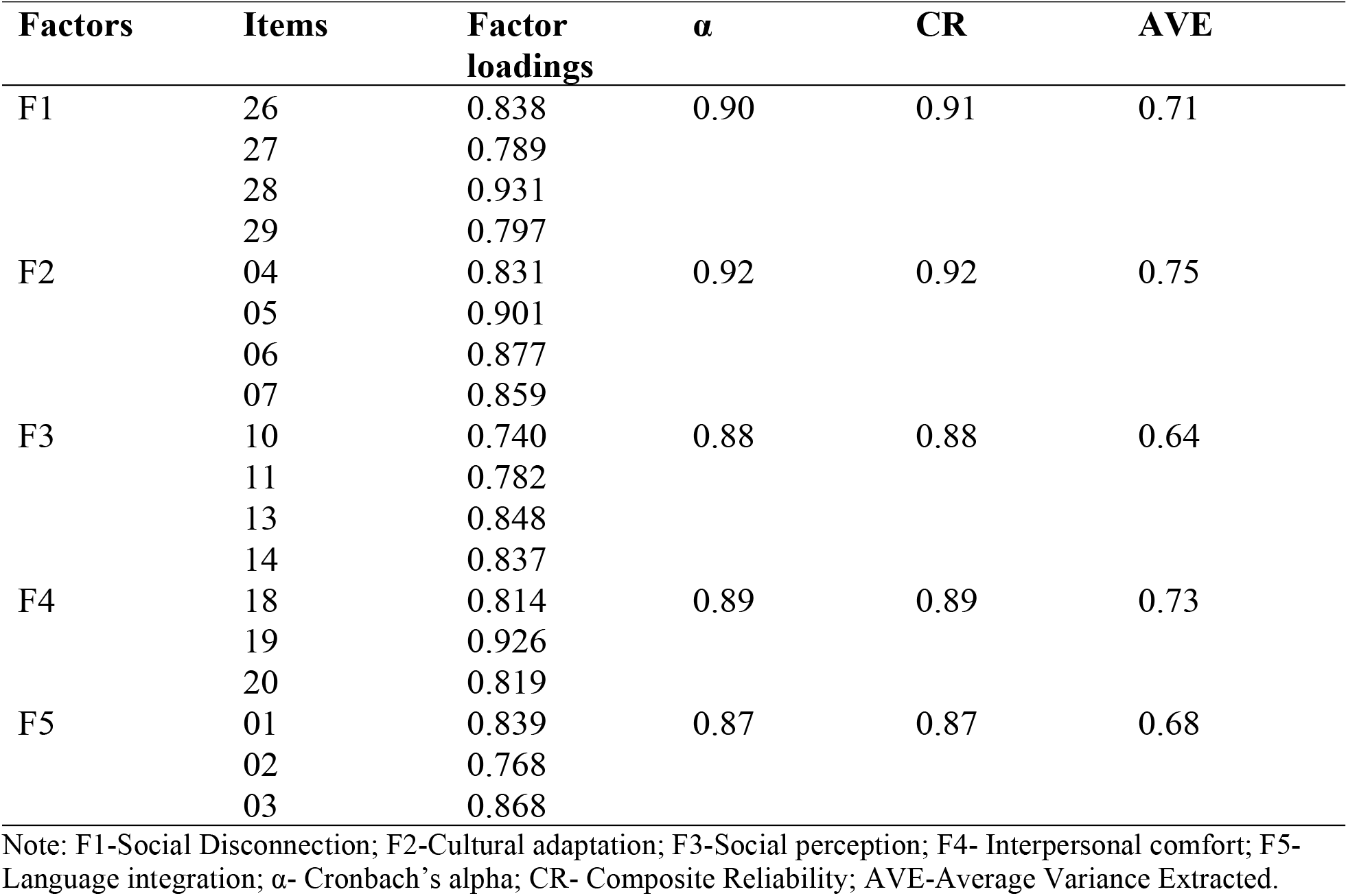
Confirmatory Structure of the Five-Dimensional Model.

| Factors | Items | Factor loadings | $\alpha$ | CR | AVE |
| --- | --- | --- | --- | --- | --- |
| F1 | 26 | 0.838 | 0.90 | 0.91 | 0.71 |
|  | 27 | 0.789 |  |  |  |
|  | 28 | 0.931 |  |  |  |
|  | 29 | 0.797 |  |  |  |
| F2 | 04 | 0.831 | 0.92 | 0.92 | 0.75 |
|  | 05 | 0.901 |  |  |  |
|  | 06 | 0.877 |  |  |  |
|  | 07 | 0.859 |  |  |  |
| F3 | 10 | 0.740 | 0.88 | 0.88 | 0.64 |
|  | 11 | 0.782 |  |  |  |
|  | 13 | 0.848 |  |  |  |
|  | 14 | 0.837 |  |  |  |
| F4 | 18 | 0.814 | 0.89 | 0.89 | 0.73 |
|  | 19 | 0.926 |  |  |  |
|  | 20 | 0.819 |  |  |  |
| F5 | 01 | 0.839 | 0.87 | 0.87 | 0.68 |
|  | 02 | 0.768 |  |  |  |
|  | 03 | 0.868 |  |  |  |

Note: F1-Social Disconnection; F2-Cultural adaptation; F3-Social perception; F4- Interpersonal comfort; F5- Language integration; $\alpha$ - Cronbach's alpha; CR- Composite Reliability; AVE-Average Variance Extracted.

**Table 4.** Discriminant validity of the SAAS.

| HTMT | F1 | F2 | F3 | F4 | F5 |
| --- | --- | --- | --- | --- | --- |
| **F1** |  |  |  |  |  |
| **F2** | 0.241 |  |  |  |  |
| **F3** | 0.376 | 0.250 |  |  |  |
| **F4** | 0.234 | 0.285 | 0.351 |  |  |
| **F5** | 0.173 | 0.065 | 0.372 | 0.342 |  |

Note: HTMT- Hetrotrait-Monotrait Ratio; F1-Social Disconnection; F2-Cultural adaptation; F3-Social perception; F4- Interpersonal comfort; F5- Language integration

The first factor, social disconnection, comprised four items demonstrating strong loadings, indicating that the factor reliably captured the underlying construct. Similarly, Cultural adaptation included four items, all of which showed excellent reliability, confirming the robustness of this factor in representing the adaptation process.

Social perception was also identified as a factor consisting of four items, with results showing good reliability and construct validity. This factor effectively measures individuals’ perceptions within social contexts.

Interpersonal comfort emerged as a factor comprising three items, exhibiting strong internal consistency and validating it as a reliable measure of comfort in interpersonal interactions.

Lastly, language integration comprises three items, each demonstrating high reliability and confirming the factor’s ability to represent language skill integration accurately.

The results indicate that all five factors demonstrate strong internal consistency and reliability. These findings suggest that the factors are valid representations of their respective constructs, capturing a substantial portion of the variance in the items. This comprehensive evaluation reaffirms the scale’s reliability and validity, providing security for measuring these specific psychological constructs.

Discriminant validity was assessed using the Heterotrait-Monotrait ratio (HTMT) to ensure that each construct within the scale is distinct from the others (see Table 4). The analysis revealed that all HTMT values were well below the conservative threshold of 0.85, typically recommended for establishing discriminant validity.

The results confirm that the constructs measured by the scale are sufficiently distinct, demonstrating strong discriminant validity. This suggests that the scale effectively captures unique dimensions of the constructs under investigation, ensuring that each factor is not merely a reflection of the others but represents a separate and meaningful concept within the broader framework of the study.

A bivariate correlation analysis assessed the relationship between the subscales of the newly validated scale and acculturative stress, focusing on the expected negative correlations (see Table 5). The Cultural Adaptation subscale showed significant negative correlations with acculturative stress factors, supporting the expected inverse relationship and indicating convergent validity. However, other subscales, such as Social Disconnection and Language Integration, showed weak positive correlations, contrary to the hypothesised negative relationship. These results suggest that while Cultural Adaptation aligns with theoretical expectations, the other subscales do not consistently demonstrate the anticipated negative relationship with acculturative stress, indicating that further investigation may be needed.

**Table 5.**
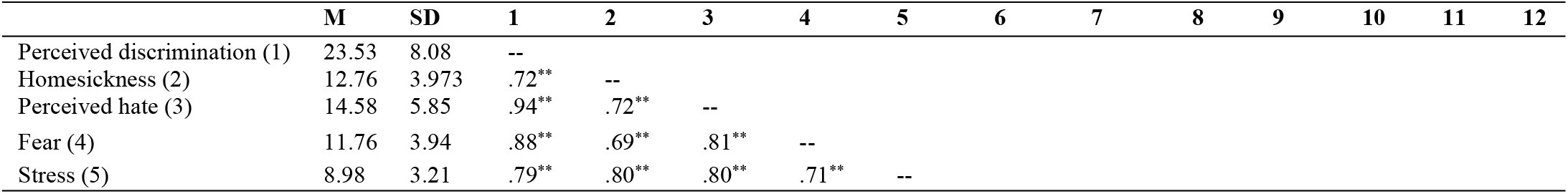
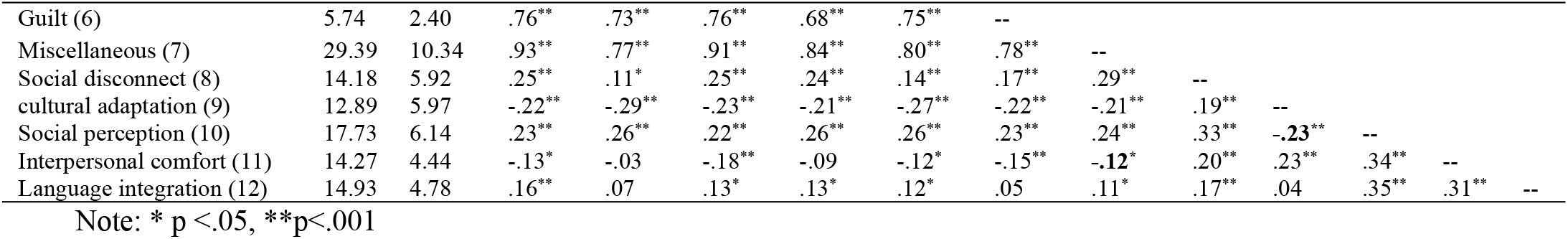
Convergent between the subscales of SAAS and acculturative stress.

|  | M | SD | 1 | 2 | 3 | 4 | 5 | 6 | 7 | 8 | 9 | 10 | 11 | 12 |
| --- | --- | --- | --- | --- | --- | --- | --- | --- | --- | --- | --- | --- | --- | --- |
| Perceived discrimination (1) | 23.53 | 8.08 | -- |  |  |  |  |  |  |  |  |  |  |  |
| Homesickness (2) | 12.76 | 3.973 | .72** | -- |  |  |  |  |  |  |  |  |  |  |
| Perceived hate (3) | 14.58 | 5.85 | .94** | .72** | -- |  |  |  |  |  |  |  |  |  |
| Fear (4) | 11.76 | 3.94 | .88** | .69** | .81** | -- |  |  |  |  |  |  |  |  |
| Stress (5) | 8.98 | 3.21 | .79** | .80** | .80** | .71** | -- |  |  |  |  |  |  |  |
| Guilt (6) | 5.74 | 2.40 | .76** | .73** | .76** | .68** | .75** | -- |  |  |  |  |  |  |
| Miscellaneous (7) | 29.39 | 10.34 | .93** | .77** | .91** | .84** | .80** | .78** | -- |  |  |  |  |  |
| Social disconnect (8) | 14.18 | 5.92 | .25** | .11* | .25** | .24** | .14** | .17** | .29** | -- |  |  |  |  |
| cultural adaptation (9) | 12.89 | 5.97 | -.22** | -.29** | -.23** | -.21** | -.27** | -.22** | -.21** | .19** | -- |  |  |  |
| Social perception (10) | 17.73 | 6.14 | .23** | .26** | .22** | .26** | .26** | .23** | .24** | .33** | -.23** | -- |  |  |
| Interpersonal comfort (11) | 14.27 | 4.44 | -.13* | -.03 | -.18** | -.09 | -.12* | -.15** | -.12* | .20** | .23** | .34** | -- |  |
| Language integration (12) | 14.93 | 4.78 | .16** | .07 | .13* | .13* | .12* | .05 | .11* | .17** | .04 | .35** | .31** | -- |

Note: \* p <.05, \*\*p<.001

### Measurement invariance

A series of nested models were tested to evaluate the scale’s measurement invariance across genders (see Table 6). The Configural invariance (Model 1), serving as the baseline model, yielded a chi-square value of χ^2^(260) = 638.95, Comparative Fit Index (CFI) = 0.92, Gamma Hat = 0.91, and McDonald’s Non-Centrality Index (Mc NCI) = 0.63. These results indicate a good fit, suggesting that the same factor structure is valid across both male and female groups. Metric invariance (Model 2) was tested by constraining the factor loadings to be equal across genders. The fit indices for this model were χ^2^(273) = 661.29, CFI = 0.92, Gamma Hat = 0.90, and Mc NCI = 0.62. The chi-square difference test comparing the configural and metric models yielded Δχ^2^(13) = 22.34, p = 0.06. Although the chi-square difference approached significance, the changes in CFI (ΔCFI = 0.002), Gamma Hat (ΔGamma Hat = 0.01), and Mc NCI (ΔMc NCI = 0.01) were minimal, supporting metric invariance. This indicates that the factor loadings are equivalent across genders, meaning the construct has the same meaning for both men and women.

**Table 6.**
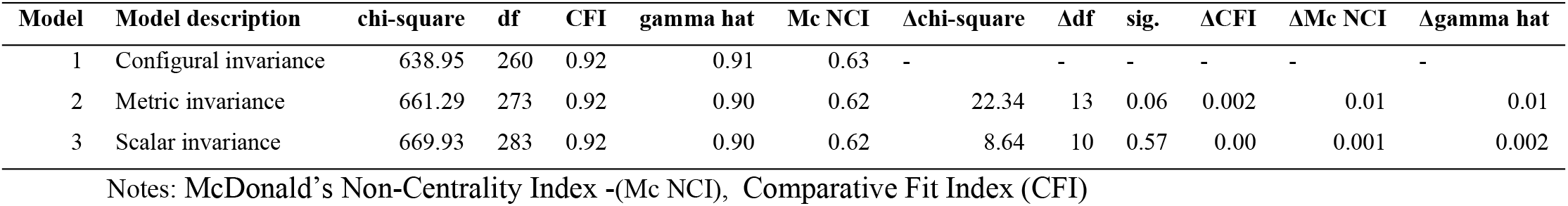
Results of tests for measurement invariance across genders.

Scalar invariance (Model 3) was assessed by further constraining the item intercepts to be equal across genders. This model produced a chi-square value of χ^2^(283) = 669.93, CFI = 0.92, Gamma Hat = 0.90, and Mc NCI = 0.62. The chi-square difference between the metric and scalar models was Δχ^2^(10) = 8.64, p = 0.57, which was not statistically significant. Additionally, changes in CFI (ΔCFI = 0.00), Gamma Hat (ΔGamma Hat = 0.002), and Mc NCI (ΔMc NCI = 0.001) were negligible, indicating that scalar invariance is supported. This suggests that the item intercepts are consistent across genders, allowing for comparing latent means between men and women.

The results indicate that the scale demonstrates configural, metric, and scalar invariance across gender. This implies that the factor structure, factor loadings, and item intercepts are consistent for both male and female groups, making meaningful comparisons of latent means between genders. Although the chi-square difference for the metric invariance test was close to significance (p = 0.06), the minimal changes in other fit indices (CFI, Gamma Hat, and Mc NCI) suggest that metric invariance holds. The support for scalar invariance confirms that differences in latent means between genders can be interpreted as true differences rather than measurement artefacts.

An independent samples t-test was conducted to explore gender differences across five psychological factors: Social Disconnection, Cultural Adaptation, Social Perception, Interpersonal Comfort, and Language Integration (see Table 7). The analysis revealed no significant differences between males and females on any of the factors. Both genders exhibited similar scores for each factor, indicating that gender does not significantly influence these constructs in this sample. This suggests that the psychological traits measured by these factors are consistent across genders.

**Table 7.**
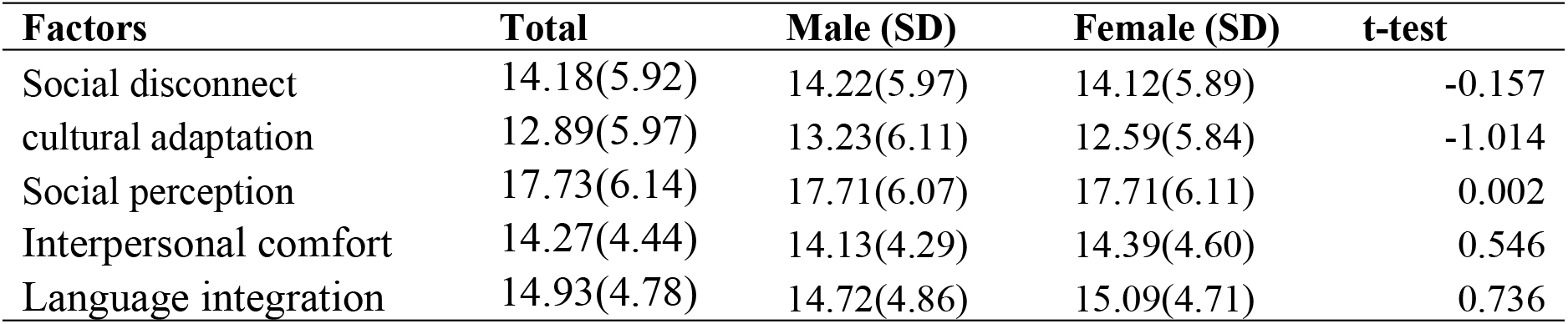
Examining latent mean differences between gender.

| Factors | Total | Male (SD) | Female (SD) | t-test |
| --- | --- | --- | --- | --- |
| Social disconnect | 14.18(5.92) | 14.22(5.97) | 14.12(5.89) | -0.157 |
| cultural adaptation | 12.89(5.97) | 13.23(6.11) | 12.59(5.84) | -1.014 |
| Social perception | 17.73(6.14) | 17.71(6.07) | 17.71(6.11) | 0.002 |
| Interpersonal comfort | 14.27(4.44) | 14.13(4.29) | 14.39(4.60) | 0.546 |
| Language integration | 14.93(4.78) | 14.72(4.86) | 15.09(4.71) | 0.736 |

## Discussion

The present study sought to adapt and validate the East Asian Acculturation Measure (EAAM) for broader applicability across diverse cultural contexts, specifically targeting populations outside the original East Asian focus. This was achieved through psychometric evaluations, including Confirmatory Factor Analysis (CFA), Exploratory Factor Analysis (EFA), and assessments of measurement invariance across gender. The findings of this study provide critical insights into the effectiveness and limitations of the adapted acculturation measure, now termed the Shortened Adapted Acculturation Scale (SAAS).

The initial CFA results indicated that the original four-factor model of the EAAM did not adequately fit the data when applied to a diverse sample comprising 87 nationalities. Despite some fit indices (e.g., SRMR, NFI, and CFI) falling within acceptable ranges, the RMSEA exceeded the commonly accepted threshold, signalling a misfit in the model’s structure. This finding underscores the challenges of applying a culturally specific measure, designed initially for East Asian populations, to a broader and more diverse population. These results align with previous research, highlighting the importance of culturally adapting psychological measures to ensure validity and reliability across different cultural groups (Demes & Geeraert, 2014).

To address the limitations observed in the CFA, an EFA was conducted, which revealed a five-factor structure that better represented the acculturation processes across the diverse sample. This new structure consisted of five distinct factors: Social Disconnection, Cultural Adaptation, Social Perception, Interpersonal Comfort, and Language Integration. The emergence of these factors reflects the complex nature of acculturation in multicultural contexts, where individuals navigate various social and cultural dynamics that the original four-factor model may not fully capture.

The five factors identified in the EFA collectively explained 51.88% of the variance, with strong loadings on each factor, indicating that the new structure provides a more accurate and comprehensive measurement of acculturation across diverse cultural groups. This finding supports the notion that acculturation is a multifaceted process that varies significantly across cultural contexts (Khawaja et al., 2014). Social Disconnection and Language Integration factors, in particular, highlight individuals’ unique challenges as they attempt to reconcile their cultural identities with those of the host culture. This theme has been noted in other studies focusing on acculturation and adaptation (Vedder et al., 2007).

The internal consistency indices for the five factors were all strong, suggesting that the adapted scale reliably measures distinct aspects of the acculturation process. The high reliability of factors such as Social Disconnection and Cultural Adaptation indicates that these dimensions are robust constructs within the broader acculturation framework. The discriminant validity analysis further supported the scale’s validity, which showed that the constructs measured by the SAAS are sufficiently distinct from one another. This is a crucial finding, as it ensures that the scale captures the unique dimensions of acculturation without conflating them with related but separate constructs.

The multiple-group CFA conducted to assess measurement invariance across genders provided additional evidence for the robustness of the adapted scale. The results indicated that the SAAS demonstrated configural, metric, and scalar invariance across male and female participants. This suggests that the scale’s factor structure, factor loadings, and item intercepts are consistent across genders, making meaningful comparisons of acculturation experiences between men and women. The support for measurement invariance is significant in ensuring that the scale can be used reliably in diverse populations without introducing gender bias. Cross-cultural research often raises this concern (Cheung & Rensvold, 2002).

### Implications for Research and Practice

The successful adaptation and validation of the EAAM into the SAAS have significant implications for research and practice. For researchers, the SAAS provides a robust tool for assessing acculturation across diverse populations, enabling more accurate and culturally sensitive studies of how individuals navigate the complexities of cultural adaptation. The scale’s ability to measure distinct aspects of acculturation, such as social disconnection and language integration, offers valuable insights into the specific challenges individuals from different cultural backgrounds face.

For practitioners, particularly those working in multicultural settings, the SAAS can inform the development of interventions to support individuals in their acculturation process. By identifying specific areas where individuals may struggle, such as social disconnection or cultural adaptation, practitioners can tailor interventions to address these challenges, thereby enhancing the effectiveness of their support.

### Limitations and Future Directions

While this study presents significant advancements in adapting and validating the EAAM for broader cultural contexts, several limitations should be noted. First, although the sample was diverse and representative of the target population, the focus on university students may limit the generalizability of the findings to other demographic groups. Acculturation processes vary significantly across age groups, socio-economic statuses, and educational levels. Therefore, future research should aim to validate the SAAS across a broader range of populations, including older adults, working professionals, and individuals with varying educational backgrounds, to capture the full spectrum of acculturation experiences.

Secondly, although the SAAS demonstrated strong psychometric properties, the study primarily focused on the scale’s structural validity without extensive exploration of its predictive validity. While the structural validity offers a solid foundation, future research should investigate how well the SAAS predicts relevant psychological and social outcomes, such as mental health and overall well-being. Understanding these relationships will provide deeper insights into the scale’s practical utility and application in diverse settings.

Finally, while the study successfully established measurement invariance across gender, other potentially relevant demographic variables, such as ethnicity, socioeconomic status, and migration history, were not examined. These factors could significantly influence acculturation experiences and should be considered in future research. Expanding the analysis to include these variables will help ensure the SAAS is robust and applicable across various subgroups within larger populations.

## Conclusion

In conclusion, the adaptation and validation of the EAAM into the SAAS represents a significant advancement in the measurement of acculturation across diverse cultural contexts. The study’s findings highlight the importance of culturally adapting psychological measures to ensure validity and reliability in multicultural settings. The SAAS offers researchers and practitioners a robust tool for assessing acculturation, providing insights into the complexities of cultural adaptation. As globalisation continues to increase cultural diversity worldwide, the need for such tools will only become more pressing, making the contributions of this study timely and essential.

## Data Availability

All relevant data are within the manuscript and its Supporting Information files.

## Acknowledgements

The authors acknowledge the support of Friedrich-Schiller University and North-West University’s financial contribution in funding this study.

